# Association of Physical Activity from Wearable Devices and Chronic Disease Risk: Insights from the All of Us Research Program

**DOI:** 10.1101/2024.11.11.24317124

**Authors:** Yu Hou, Erjia Cui, Sayeed Ikramuddin, Rui Zhang

**Affiliations:** Division of Computational Health Sciences, University of Minnesota, Minneapolis, Minnesota, USA; Division of Biostatistics & Health Data Science, University of Minnesota, Minneapolis, Minnesota, USA; Department of Surgery, University of Minnesota, Minneapolis, Minnesota, USA

**Keywords:** Physical activity, Fitbit, Chronic disease, Wearable devices, All of Us Research Program

## Abstract

**Background:** Physical activity is widely recognized as a key modifiable factor for reducing the risk of chronic diseases. Wearable devices such as Fitbit offer a unique opportunity to objectively measure physical activity metrics, providing insights into the association between different types of physical activity and chronic disease risk.

**Objective:** This study aims to examine the association between physical activity metrics derived from Fitbit devices and the incidence of various chronic diseases among participants from the All of Us Research Program.

**Methods:** Physical activity metrics included daily steps, elevation gain, and activity durations at different intensities (e.g., very active, lightly active, and sedentary). Cox proportional hazards models and multiple regression models were used to assess the relationship between these metrics and the incidence of chronic diseases represented by Phecodes. Age, sex, and body mass index (BMI) were included as covariates.

**Results:** A total of 15,538 participants provided Fitbit activity data, of which 9,320 also had electronic health records (EHR). Increased daily step count, elevation gain, and very active minutes were significantly associated with a reduced risk of several chronic conditions, including obesity, Type 2 diabetes, and major depressive disorder. Conversely, increased sedentary time was linked to higher risks for conditions such as obesity, Type 2 diabetes, and essential hypertension. Multiple regression analyses confirmed these associations.

**Conclusion:** Our findings highlight the beneficial effects of increased physical activity, particularly daily steps and elevation gain, on reducing the risk of chronic diseases. Conversely, sedentary behavior remains a significant risk factor for the development of several conditions. These insights may inform personalized activity recommendations aimed at reducing disease burden and improving population health outcomes.

## Introduction

Chronic diseases, such as cardiometabolic disorders, obesity, and major depressive disorder (MDD), are the main causes of morbidity and mortality worldwide^1–3^. As lifestyle factors play a pivotal role in their development and progression, increasing attention has been given to the potential of physical activity as a modifiable factor^4,5^. Wearable devices offer a unique opportunity to non-invasively capture continuous, objective measures of physical activity, providing the chance to investigate activity patterns and their associations with chronic disease outcomes. Previous studies have highlighted the association between physical activity and various chronic diseases, including cardiovascular disease, type 2 diabetes, depression, Parkinson’s disease, and Multiple Sclerosis^4–8^. However, most of these studies have relied on either self-reported physical activity questionnaires or shorter periods of objective data collection using devices like accelerometers or pedometers. While self-reported data provide general estimates, they often deviate from directly measured physical activity levels^9,10^.

With the advent of commercial wearable devices, such as Fitbit, there is now an opportunity to overcome many of these challenges in physical activity research by gathering long-term, objective activity data. Studies using Fitbit devices have demonstrated their validity and reliability in measuring activity metrics such as steps, heart rate, and energy expenditure^11,12^. The longitudinal data collected by these devices offer unprecedented insight into individual activity patterns, making it feasible to conduct large-scale epidemiological studies to investigate associations between real-world activity behaviors and chronic disease development.

The All of Us Research Program (AoU), a National Institutes of Health-funded initiative, aims to collect health data from over one million individuals across the United States, representing a diverse range of populations^13,14^. This program integrates multiple sources of health-related information, including electronic health records (EHRs), genomic data, physical measurements, and participant surveys. A subset of participants who own Fitbit devices has been invited to voluntarily share their Fitbit data, including activity and sleep patterns, with a median monitoring time of 4.5 years. This unique dataset enables, for the first time, the exploration of longitudinal associations between objectively measured activity behaviors and multiple chronic disease outcomes in a large, diverse cohort of the United States.

In this study, we leveraged the extensive Fitbit activity data and longitudinal EHRs available through the AoU to investigate the associations between real-world physical activity patterns and chronic disease incidence. Our study aims to provide a comprehensive understanding of how daily activity, in terms of intensity, duration, and regularity, may influence the risk of developing chronic diseases, with findings that may contribute to evidence-based lifestyle recommendations for disease prevention in the general US population.

## Methods

### Study Participants

All study participants consented to participate in the AoU, a large-scale initiative funded by the National Institutes of Health aimed at accelerating research in health and disease^13,14^. All adults (aged ≥18 years) living in the United States were eligible to enroll, except individuals who were incarcerated or unable to provide consent independently. For this study, we used the registered and controlled tier data (C2022Q4R9) available on the AoU Researcher Workbench. Data from 413,457 individuals who were enrolled from May 2018 to July 1, 2022, were available at the time of analysis^15^. The data collection included detailed demographic information and survey responses gathered during digital enrollment, physical measurements and vital signs recorded by partnered healthcare provider organizations, voluntarily shared EHRs from these partners, and Fitbit data shared by participants through linkage of their Google Fitbit account within the AoU participant portal^13,14,16^.

### Fitbit activity data

For this study, we used Fitbit-derived daily activity data that included various features providing detailed insights into the physical activity pattern of participants. The features include activity calories, calories burned from basal metabolic rate (BMR), total calories burned, elevation traveled, minutes of activity at different intensity levels, and total number of steps. Fitbit activity data offers a unique advantage over traditional self-reported activity measures due to its objective, continuous, and non-invasive tracking. The availability of detailed activity metrics, such as different activity intensities (e.g., light, moderate, and vigorous), provides a comprehensive understanding of participants’ activity profiles over time. Specifically, the Fitbit data was collected from participants’ Fitbit devices after linking their accounts through the All of Us portal, allowing us to capture longitudinal activity behaviors.

In this study, we focused on summarizing daily activity metrics for each participant across their monitoring period by averaging the daily activity patterns to determine a typical activity profile for each participant. This allowed us to characterize individual variability and better understand the association between activity behaviors and health outcomes. Moreover, the metrics allowed us to capture not just overall activity but also to differentiate between varying intensities of activity, helping to provide insights into both total activity volume and how activity was distributed throughout the day.

### Preprocessing

The preprocessing of Fitbit activity data involved multiple steps to ensure a clean and reliable cohort for analysis. The dataset was accessed using Google BigQuery, where the initial data was extracted including all participants with available Fitbit data. The activity data features were extracted for individuals with activity summary records using SQL queries, which included metrics such as activity calories, BMR, calories out, elevation, minutes of activity at light, moderate, and vigorous levels, and total steps.

To prepare the data for analysis, we followed a cohort filtering process to ensure participants met key inclusion criteria (supplement Fig. 1). First, participants were filtered to include only those aged 18 years or older. The age of each participant was calculated by comparing the date of their first recorded Fitbit activity with their date of birth and only those who were adults at the time of their first activity were retained. Next, participants were required to have more than three days of Fitbit activity records. This filter was commonly used in the literature^17,18^ to ensure sufficient data quality and to exclude participants with limited activity records, which could lead to unreliable results. Additionally, participants were included only if they had available linked EHR data. This ensured that all participants had corresponding EHR information, enabling the assessment of associations between Fitbit-derived activity metrics and clinical outcomes.

### Outcomes and covariates

In this study, the outcomes were derived from diagnostic events recorded in the participants’ EHR. Diagnostic events were identified using International Classification of Diseases (ICD) codes, including both ICD-9 and ICD-10 versions. These ICD codes were mapped to 1,759 distinct phecodes using the PheWAS catalog’s Phecode Maps^19–21^, which allowed for consistent categorization of diagnoses. Each phecode was also linked to its corresponding phenotype and category. Phecodes with fewer than 20 cases in the dataset were excluded to ensure statistical robustness. Covariates for the analysis were extracted from the demographics data and included age and sex.

### Statistical analyses

We used multiple logistic regression models to assess the association between physical activity metrics (e.g., steps, activity calories, and minutes at different activity intensities) and health outcomes. The outcomes in this study were the presence or absence of specific diagnoses, represented by phecodes, which were derived from EHR. To ensure robustness, participants who had a diagnosis event for a given phecode within the initial 180 days of their Fitbit monitoring period were excluded. Analyses were adjusted for age, sex, and body mass index (BMI) at baseline. Bonferroni correction was applied to adjust for multiple comparisons.

We further performed Cox proportional hazards regression models to explore the association between physical activity metrics and selected chronic diseases. These chronic diseases were specifically chosen based on associations reported in previous studies, allowing us to focus on conditions with established links to physical activity^22–29^. The primary predictor variables were derived from Fitbit activity data, which included activity metrics such as activity calories, elevation, minutes of activity at different intensities (e.g., fairly active, lightly active, very active), and steps. The outcomes were defined as diagnosis events from EHR, represented by phecodes, which captured specific health conditions. For each phecode, we defined the baseline as the average of activity metrics during the initial 180 days after each participant’s first Fitbit activity record. Individuals who had fewer than 15 days of recorded activity during this baseline period were excluded to ensure data reliability. Additionally, participants who had been diagnosed before or within the initial 180 days of the Fitbit activity record were excluded from the analysis for that particular phecode to mitigate potential reverse causation. The Cox model was fit by including the activity metric as a predictor one at a time, adjusting for age, sex and BMI. The event variable was set to indicate whether a diagnosis occurred during the follow-up period, while the duration variable indicated the number of days from the start of the observation period until the diagnosis or the end of the follow-up. For the activity metrics, daily steps were analyzed in increments of 2,000 steps to explore the effect of every 2,000-step increase on disease risk. Elevation gain was analyzed in increments of 100 meters, allowing us to assess the impact of every additional 100 meters on disease risk. Furthermore, the durations of various activity types (e.g., lightly active, very active) as well as sedentary time were converted to hours to evaluate the effect of each additional hour on disease risk.

Statistical analyses were performed using Python (v3.8, https://www.python.org/) on the AoU Researcher Workbench, a secure cloud-based platform. The analyses were conducted by the libraries including Pandas, NumPy, Statsmodels, and Lifelines.

## Results

A total of 15,538 participants from the AoU Research Program had available Fitbit activity data. Among these, 9,320 participants also had corresponding EHR data. The median age of participants with Fitbit activity data was 51 years (interquartile range: 36 to 62 years). Most participants were female (69.33%), while 27.92% were male, and 2.75% had unknown sex information. Regarding race, 81.89% identified as White, 5.30% as Black, 2.85% as Asian, 2.98% as Other, and 6.97% had unknown race information. A small proportion (5.69%) identified as Hispanic or Latino, while 90.48% were not Hispanic or Latino, and 3.83% had unknown ethnicity. The median Fitbit monitoring length was 3.04 years (1.43, 5.09). The median daily step count was 7,194 steps (5,308, 9,354). Participants burned a median of 867.30 activity calories per day (640.21, 1123.62), with a median BMR of 1471.67 calories (1287.68, 1678.99). The median total daily calories burned was 2212.83 (1902.28, 2599.22). The median elevation gained was 71.84 meters (34.33, 132.20). In terms of activity intensity, the median daily sedentary minutes was 862.58 (743.44, 1054.02), lightly active minutes was 193.59 (152.04, 238.56), fairly active minutes was 13.86 (7.81, 23.25), and very active minutes was 13.09 (6.08, 25.81) (Table 1).

**Table 1.** Baseline characteristics of study participants.

| Variable | Fitbit activity participants |
| --- | --- |
| Age | 51 (36, 62) |
| Sex |  |
| Female | 6,462 (69.33%) |
| Male | 2,602 (27.92%) |
| Race |  |
| White | 7,632 (81.9%) |
| Black | 494 (5.30%) |
| Asian | 266 (2.85%) |
| Others | 278 (2.98%) |
| Unknown | 650 (6.97%) |
| Ethnicity |  |
| Hispanic or Latino | 530 (5.69%) |
| Non-Hispanic or Latino | 8,433 (90.48%) |
| Unknown | 357 (3.83%) |
| Fitbit monitoring length | 3.04 (1.43, 5.09) years |
| Daily steps | 7,194 (5,308, 9,354) |
| Daily elevation | 74.84 (34.33, 132.20) meters |
| Activity intensity (minutes) |  |
| Lightly active | 193.59 (152.04, 238.56) |
| Very active | 13.09 (6.08, 25.81) |
| Fairly active | 13.86 (7.81, 23.25) |
| Sedentary | 862.58 (743.44, 1054.02) |
| Calories |  |
| Daily activity calories <sup>a</sup> | 867.30 (640.21, 1123.62) |
| Daily BMR calories <sup>b</sup> | 1471.67 (1287.68, 1678.99) |
| Daily calories burned <sup>c</sup> | 2212.83 (1902.28, 2599.22) |
| <sup>a</sup> The number of calories burned for the day during periods the user was active above the sedentary level. This includes both activities burned calories and BMR.<br><sup>b</sup> Total BMR calories burned for the day.<br><sup>c</sup> Total calories burned for the day |  |

### Associations between physical activity patterns and chronic disease incidence

A total of 31 significant associations with diseases were identified after Bonferroni correction through the exploration of associations between physical activity metrics and health outcomes (Fig. 1). Overall, daily step count was significantly associated with 12 diseases, while activity calories showed a significant association with 1 disease. Nine diseases were significantly related to BMR calories burned, and three were linked to total daily calories burned. The elevation gained (height climbed daily) was associated with 15 diseases. Additionally, 1 disease was significantly associated with sedentary minutes, while lightly active minutes were linked to 1 disease, fairly active minutes to 4 diseases, and very active minutes to 4 diseases (Fig 2).

**Fig. 1.**
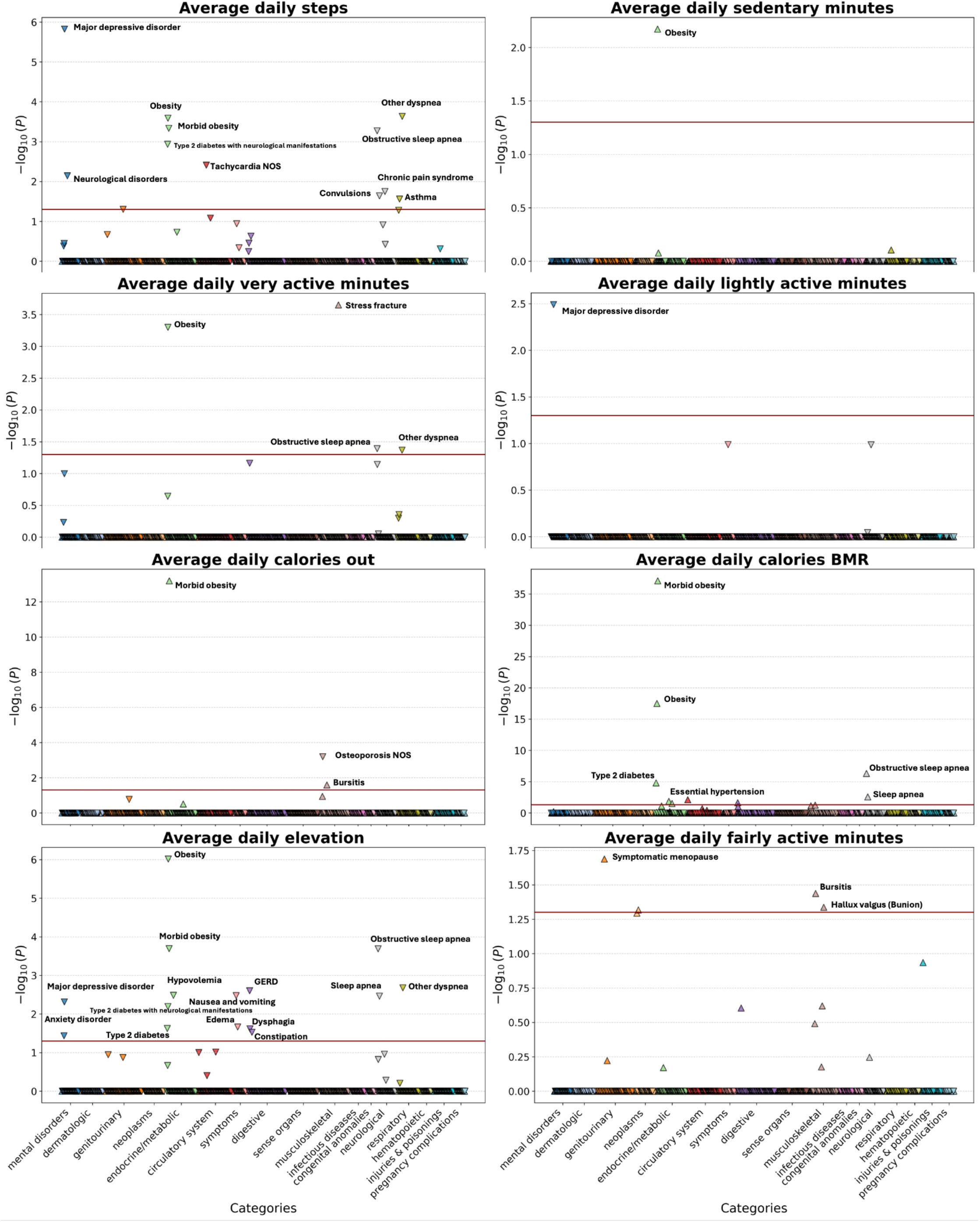
Associations between physical activity patterns and chronic disease incidence

**Fig. 2.**
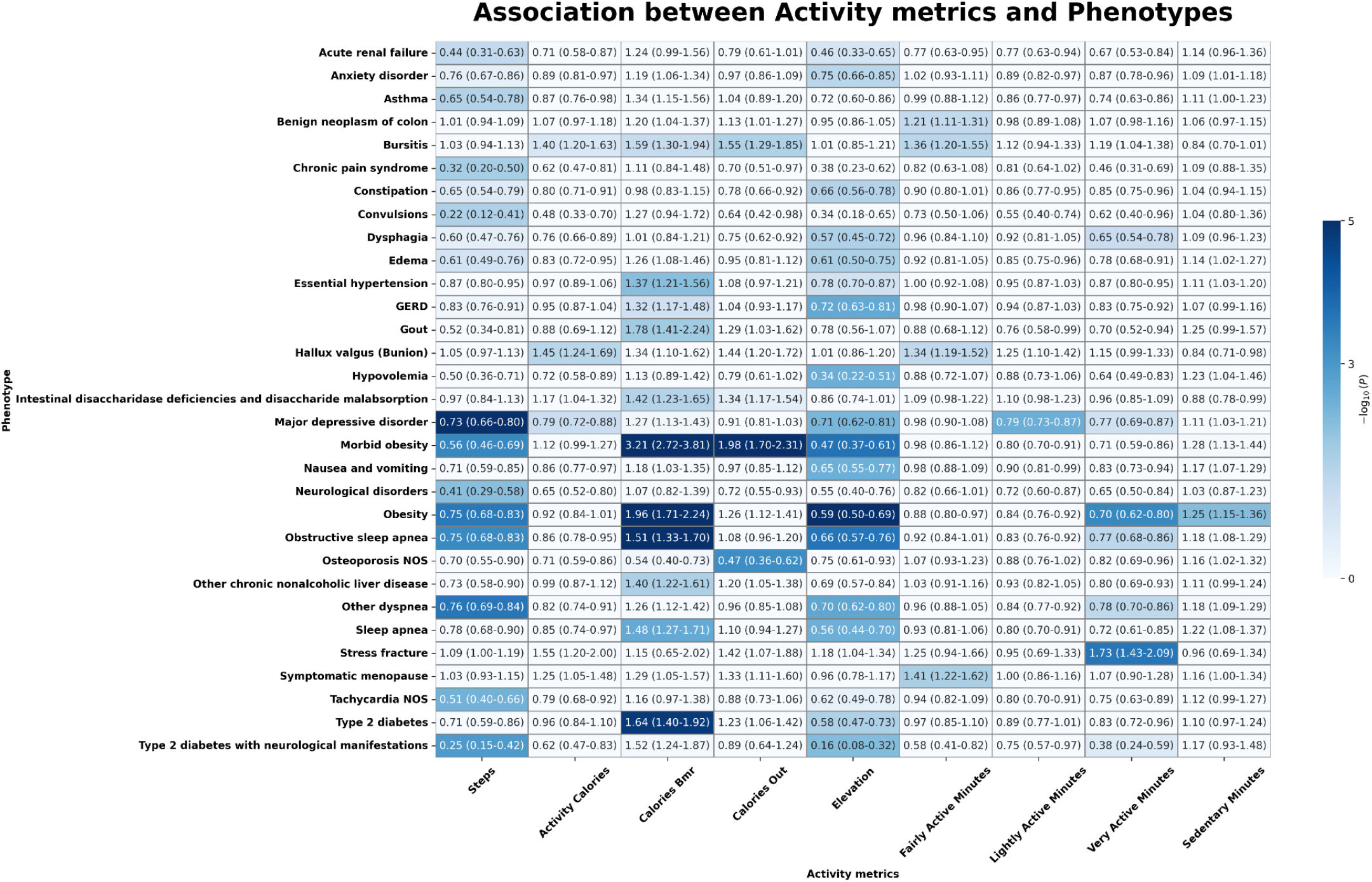
Heatmap of significant phenotypes across the physical activity patterns

The increase in daily steps was significantly associated with a reduced risk of Type 2 diabetes with neurological manifestations (odds ratio (OR) = 0.25; 95% confidence interval (CI) = 0.15– 0.42; p-value = 0.0012), obstructive sleep apnea (0.75; 0.68–0.83; 0.0005), obesity (0.75; 0.68– 0.83; 0.0003), major depressive disorder (0.73; 0.66–0.80; <0.0001), and respiratory diseases such as asthma (0.65; 0.54–0.78; 0.0274) and other dyspnea (0.76; 0.69–0.84; 0.0002). Neurological disorders, including convulsions (0.22; 0.12–0.41; 0.0230), and chronic pain syndrome (0.32; 0.20–0.50; 0.0177) were also significantly associated with increased steps. Tachycardia NOS (0.51; 0.40–0.66; 0.0039) and acute renal failure (0.44; 0.31–0.63; 0.0496) were also significantly associated with increased steps.

The increase in BMR calories burned was significantly associated with an increased risk of essential hypertension (1.37; 1.21–1.56; 0.0073), Type 2 diabetes (1.64; 1.40–1.92; 0.00002), other chronic nonalcoholic liver disease (1.40; 1.22–1.61; 0.0228), obstructive sleep apnea (1.51; 1.33–1.70; <0.0001), and obesity (1.96; 1.71–2.24; <0.0001). Morbid obesity was highly associated with increased BMR calories burned (3.21; 2.72–3.81; <0.0001), as were sleep apnea (1.48; 1.27–1.71; 0.0026), and gout (1.78; 1.41–2.24; 0.0140).

An increase in daily elevation gain was significantly associated with a reduced risk of several conditions. Specifically, higher elevation gain was significantly associated with a reduced risk of Type 2 diabetes (0.58; 0.47–0.73; 0.0236), gastroesophageal reflux disease (GERD) (0.72; 0.63– 0.81; 0.0025), Type 2 diabetes with neurological manifestations (0.16; 0.08–0.32; 0.0064), and obstructive sleep apnea (0.66; 0.57–0.76; <0.0001). Additionally, elevation gain was associated with a lower risk of nausea and vomiting (0.65; 0.55–0.77; 0.0033), obesity (0.59; 0.50–0.69; <0.0001), dysphagia (0.57; 0.45–0.72; 0.0241), and anxiety disorder (0.75; 0.66–0.85; 0.0364). Major depressive disorder was also significantly associated with increased elevation gain (0.71; 0.62–0.81; 0.0048), as were morbid obesity (0.47; 0.37–0.61; 0.0002), sleep apnea (0.56; 0.44– 0.70; 0.0034), and constipation (0.66; 0.56–0.78; 0.0296). Other conditions significantly associated with increased elevation gain included edema (0.61; 0.50–0.75; 0.0216), other dyspnea (0.70; 0.62–0.80; 0.0021), and hypovolemia (0.34; 0.22–0.51; 0.0328).

An increase in very active minutes was significantly associated with a reduced risk of obstructive sleep apnea (0.77; 0.68–0.86; 0.0406), obesity (0.70; 0.62–0.80; 0.0005), and other dyspnea (0.78; 0.70–0.86; 0.0427). However, an increase in very active minutes was also associated with an increased risk of stress fractures (1.73; 1.43–2.09; 0.0002). Fairly active minutes were significantly associated with an increased risk of bursitis (1.36; 1.20–1.55; 0.04). Lightly active minutes were significantly associated with a reduced risk of major depressive disorder (0.79; 0.73–0.87; 0.003). Lastly, an increase in sedentary minutes was significantly associated with an increased risk of obesity (1.25; 1.15–1.36; 0.007).

### Cox Model Results for Physical Activity and Chronic Diseases

The Cox proportional hazards regression models indicated several significant associations between physical activity metrics and chronic disease risk (Table 2; supplementary Fig. 1 - 8). For every 2,000-step increase in daily steps, the hazard ratio (HR) for obesity was 0.91 (95% CI: 0.87–0.95), for morbid obesity it was 0.72 (0.68–0.77), and for Type 2 diabetes it was 0.83 (0.80– 0.86). Major depressive disorder was also significantly associated with increased steps, with an HR of 0.81 (0.78–0.84). The risk of essential hypertension decreased with increasing steps, with an HR of 0.91 (0.88–0.93). Chronic pain syndrome showed an HR of 0.75 (0.66–0.85), suggesting a significant reduction in risk with increased steps.

**Table 2.** Cox proportional hazards models for the association between Fitbit activity metric and chronic diseases.

|  | Steps <sup>a</sup> | Elevation <sup>b</sup> | Sedentary <sup>c</sup> | Lightly active <sup>c</sup> | Very active <sup>c</sup> |
| --- | --- | --- | --- | --- | --- |
| Chronic disease | HR (95% CI) |  |  |  |  |
| Obesity | 0.911 (0.871-0.952) | 0.683 (0.613-0.761) | 1.056 (1.038-1.076) | 0.860 (0.812-0.910) | 0.056 (0.037-0.084) |
| Morbid obesity | 0.720 (0.676-0.766) | 0.537 (0.456-0.633) | 1.154 (1.131-1.179) | 0.935 (0.873-1.002) | 0.003 (0.002-0.008) |
| Type 2 diabetes | 0.831 (0.798-0.864) | 0.670 (0.611-0.735) | 1.042 (1.026-1.058) | 0.869 (0.827-0.914) | 0.442 (0.354-0.551) |
| Major depressive disorder | 0.806 (0.776-0.838) | 0.739 (0.681-0.802) | 1.058 (1.043-1.074) | 0.807 (0.771-0.845) | 0.324 (0.252-0.417) |
| Obstructive sleep apnea | 1.001 (0.968-1.034) | 0.808 (0.751-0.870) | 1.044 (1.027-1.061) | 0.862 (0.819-0.908) | 0.513 (0.414-0.634) |
| Other dyspnea | 0.796 (0.747-0.848) | 0.814 (0.726-0.913) | 1.048 (1.023-1.074) | 0.858 (0.794-0.928) | 0.318 (0.215-0.472) |
| Essential hypertension | 0.907 (0.884-0.929) | 0.789 (0.750-0.830) | 1.065 (1.054-1.076) | 0.888 (0.858-0.919) | 0.582 (0.510-0.664) |
| Chronic pain syndrome | 0.751 (0.661-0.854) | 0.670 (0.504-0.890) | 0.967 (0.917-1.019) | 1.243 (1.200-1.288) | 4.8e<sup>-4</sup> (4.9e<sup>-5</sup>-4.7e<sup>-3</sup>) |
| <sup>a</sup> : 2,000 steps<br><sup>b</sup> : 100 m<br><sup>c</sup> : Hours |  |  |  |  |  |

For elevation, each 100-meter increase was significantly associated with a reduced risk of several conditions, including obesity (HR = 0.68; 95% CI: 0.61–0.76), morbid obesity (0.54; 0.46–0.63), Type 2 diabetes (0.67; 0.61–0.74), and major depressive disorder (0.74; 0.68–0.80). Other conditions, such as obstructive sleep apnea (0.81; 0.75–0.87) and essential hypertension (0.79; 0.75–0.83), also showed reduced risk with increased elevation.

Sedentary time was positively associated with increased risk for multiple conditions. Specifically, for each additional hour of sedentary time, the risk of obesity increased (HR = 1.06; 95% CI: 1.04– 1.08), as did the risks of morbid obesity (1.15; 1.13–1.18), Type 2 diabetes (1.04; 1.03–1.06), and major depressive disorder (1.06; 1.04–1.07). Essential hypertension (1.07; 1.05–1.08) also showed increased risk with more sedentary time.

Lightly active time was associated with a reduced risk of several conditions. Each additional hour of light activity was linked to a decreased risk of obesity (HR = 0.86; 95% CI: 0.81–0.91), Type 2 diabetes (0.87; 0.83–0.91), major depressive disorder (0.81; 0.77–0.85), and obstructive sleep apnea (0.86; 0.82–0.91). For very active time, each additional hour was significantly associated with a reduced risk of multiple conditions, including obesity (HR = 0.06; 95% CI: 0.04–0.08), morbid obesity (0.003; 0.002–0.008), Type 2 diabetes (0.44; 0.35–0.55), and major depressive disorder (0.32; 0.25–0.42). Conversely, very active minutes were also associated with an increased risk of chronic pain syndrome (HR = 4e^-4^; 95% CI: 4.9e^-5^–4.7e^-3^), suggesting a complex relationship between high-intensity activity and certain conditions.

## Discussion

This study investigated the association between physical activity metrics, derived from Fitbit wearable devices, and the incidence of various chronic diseases, using data from the AoU Research Program. Our findings indicate that increased physical activity, represented by daily step count, elevation gain, and activity at different intensities, is generally associated with a reduced risk of several chronic diseases, including obesity, Type 2 diabetes, major depressive disorder, and obstructive sleep apnea. Conversely, increased sedentary time was associated with an elevated risk of these conditions. These findings contribute to a growing body of literature highlighting the importance of regular physical activity in promoting overall health and reducing disease risk.

The observed inverse association between daily step count and the risk of developing obesity, Type 2 diabetes, and major depressive disorder aligns with previous studies that have demonstrated the protective effect of increased daily steps on cardiometabolic health and mental well-being^5,30–32^. Specifically, the results suggest that for increased steps, there is a significant reduction in the risk of these conditions. This is consistent with the idea that increased daily physical activity improves energy expenditure, enhances metabolic health, and plays a role in mitigating depressive symptoms^4,33^. These findings support public health recommendations that encourage individuals to achieve a higher daily step count as a feasible strategy for reducing the risk of chronic disease.

Our study also found that elevation gain, a less commonly explored metric, is significantly associated with a lower risk of several chronic diseases, including Type 2 diabetes, morbid obesity, and obstructive sleep apnea. Previous research has indicated that activities involving increased elevation, such as stair climbing, may have potential health benefits, though more direct evidence is needed to confirm their effects on cardiovascular and metabolic outcomes^32,34^. This suggests that incorporating activities with an elevation component, even in everyday routines, could provide additional health benefits. Future research should further explore the role of elevation-related activities in reducing chronic disease risk.

The association between activity intensity and health outcomes was also highlighted in this study. Very active were found to significantly reduce the risk of obesity, Type 2 diabetes, and obstructive sleep apnea, whereas increased fairly active and lightly active were associated with reduced risk of several chronic conditions, including major depressive disorder. These findings align with previous research that has demonstrated the benefit of different intensities of physical activity on cardiometabolic health and mental health outcomes^35,36^. However, it is noteworthy that being very active was also linked to an increased risk of stress fractures, highlighting that high-intensity physical activity, while beneficial for preventing chronic diseases, may also pose certain risks, particularly to musculoskeletal health. This finding suggests a need for personalized physical activity recommendations, particularly for populations at risk of musculoskeletal injuries^37^.

Conversely, increased sedentary time was consistently associated with an elevated risk of multiple chronic diseases, including obesity, Type 2 diabetes, and essential hypertension. This is consistent with prior studies that have reported the detrimental effects of prolonged sedentary behavior on metabolic health and the increased risk of cardiometabolic diseases^38–40^. The significant association between sedentary behavior and these conditions underscores the importance of reducing sedentary time as a complementary strategy to increasing physical activity for improving health outcomes.

One of the key strengths of this study is the use of real-world wearable data, which provides objective and granular information on physical activity patterns over an extended period. Unlike traditional epidemiological studies that rely on self-reported physical activity, wearable-derived metrics offer greater accuracy and enable the assessment of various dimensions of physical activity, including intensity, duration, and elevation. However, there are several limitations to consider. First, the observational nature of the study precludes establishing causation. Despite efforts to mitigate reverse causation by excluding participants diagnosed within the initial 180 days, residual confounding may still exist. Additionally, the reliance on Fitbit users may introduce selection bias, as these individuals may be more health-conscious compared to the general population.

In conclusion, our findings underscore the importance of increasing daily steps, incorporating elevation-related activities, and engaging in varying intensities of physical activity to reduce the risk of chronic diseases. Reducing sedentary behavior is equally crucial in mitigating the risk of these conditions. These results provide valuable insights for developing targeted public health interventions aimed at promoting physical activity and reducing sedentary behavior, ultimately contributing to improved population health outcomes. Future studies should focus on elucidating the mechanisms underlying these associations and exploring personalized activity recommendations to maximize health benefits while minimizing risks.

## Conclusion

This study highlights the significant role that physical activity metrics, as measured by wearable devices, play in reducing the risk of chronic diseases. Higher daily step counts, increased elevation gain, and more time spent in physical activities of varying intensities are associated with a lower risk of conditions like obesity, Type 2 diabetes, and major depressive disorder, while increased sedentary time remains a significant risk factor. These findings underscore the need for public health initiatives to promote diverse physical activities and reduce sedentary behavior, ultimately contributing to improved health outcomes and reduced chronic disease burden.

## Author contributions

Y.H. and R. Z. conceived the study. Y. H. served as the principal author, extracting data, designing and conducting the entire experiment and write the manuscript. E. C. assisted in conceiving the study design and reviewed the manuscript. S. I. provided clinical expertise and reviewed the manuscript. All authors contributed to the production of the final manuscript.

## Data availability

This study used data from the All of Us Research Program’s Control Tier Dataset v7, available to authorized users on the Researcher Workbench, which can be accessed via https://workbench.researchallofus.org/.

## Code availability

Code used for this study is available to approved researchers upon request. Researchers can access the code on the All of Us Research Workbench platform by contacting our study team.

