## Supplemental Figures for "Association of Physical Activity from Wearable Devices and Chronic Disease Risk: Insights from the All of Us Research Program"

### Supplementary Figures

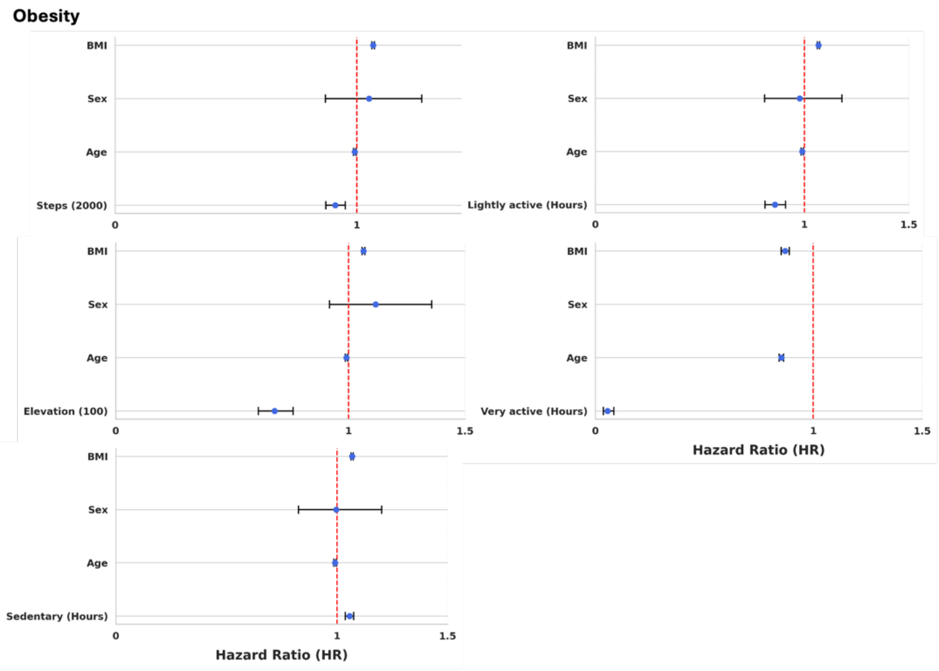


**Supplementary Fig. 1** Forest plots from Cox proportional hazard models for associations between Fitbit activity metric and obesity.

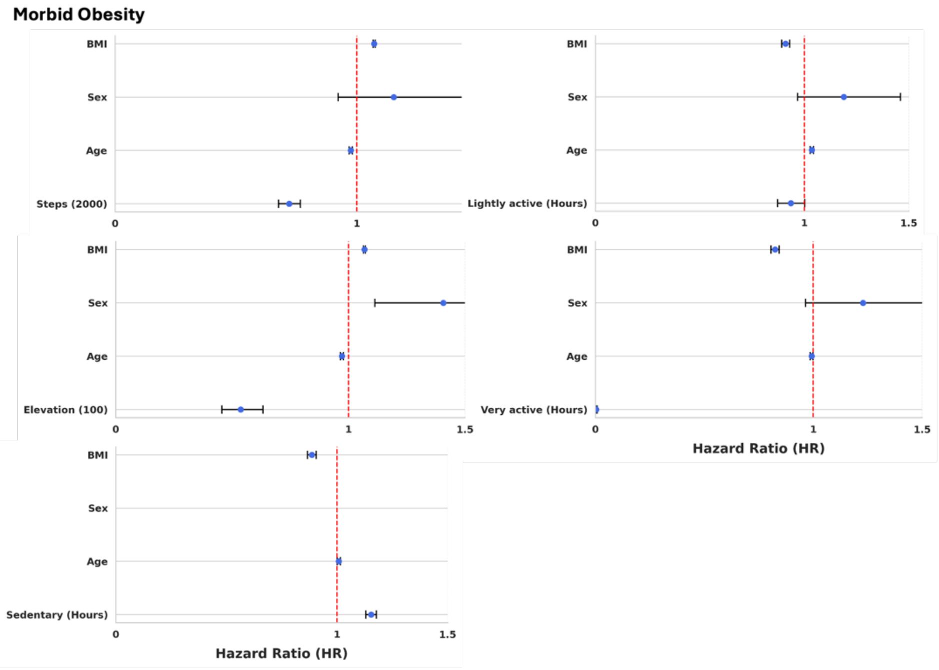


**Supplementary Fig. 2** Forest plots from Cox proportional hazard models for associations between Fitbit activity metric and morbid obesity.

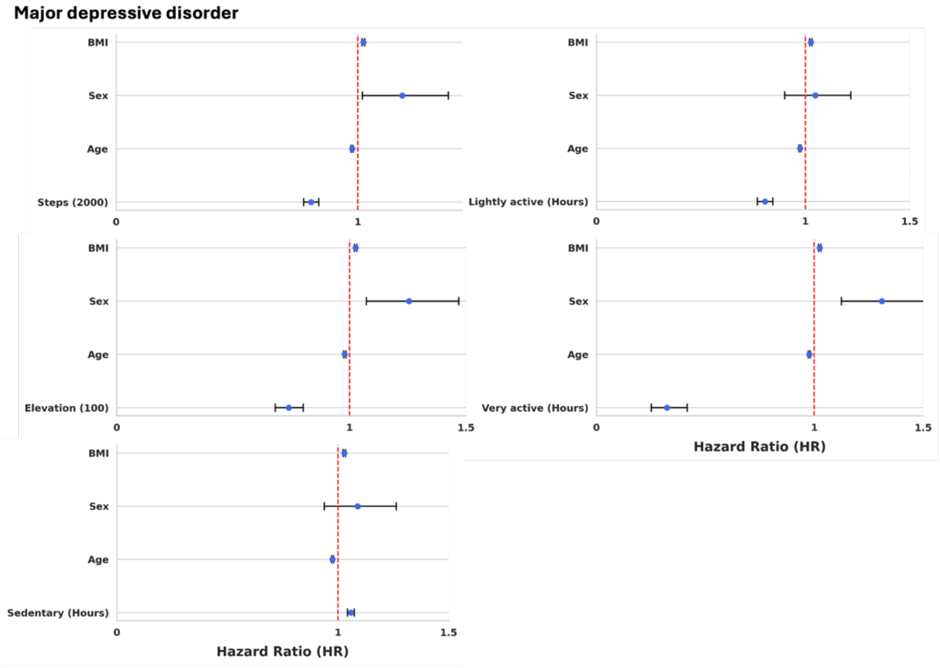


**Supplementary Fig. 3** Forest plots from Cox proportional hazard models for associations between Fitbit activity metric and major depressive disorder.

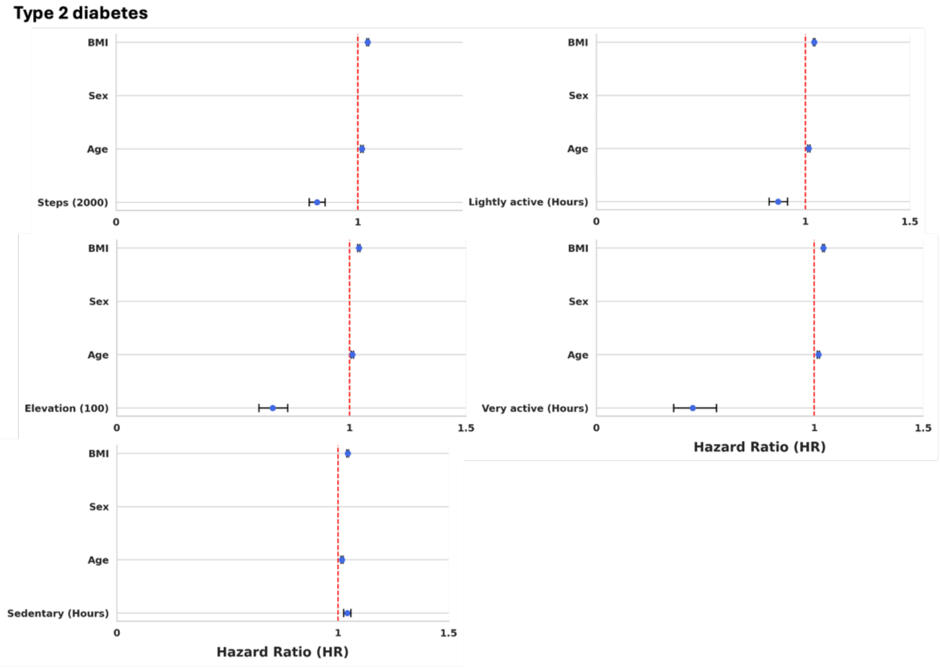


**Supplementary Fig. 4** Forest plots from Cox proportional hazard models for associations between Fitbit activity metric and Type 2 diabetes.

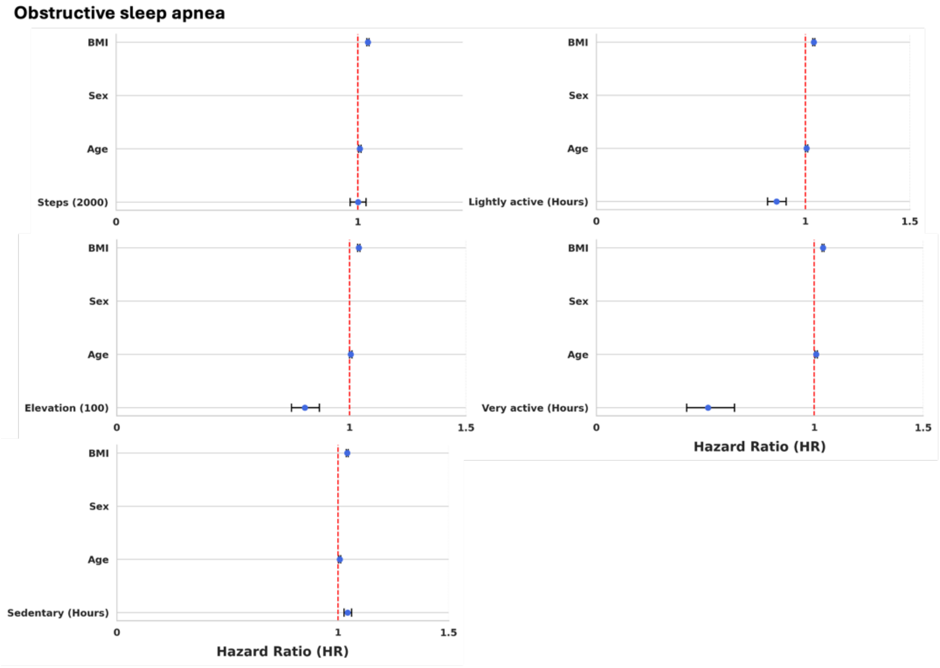


**Supplementary Fig. 5** Forest plots from Cox proportional hazard models for associations between Fitbit activity metric and obstructive sleep apnea.

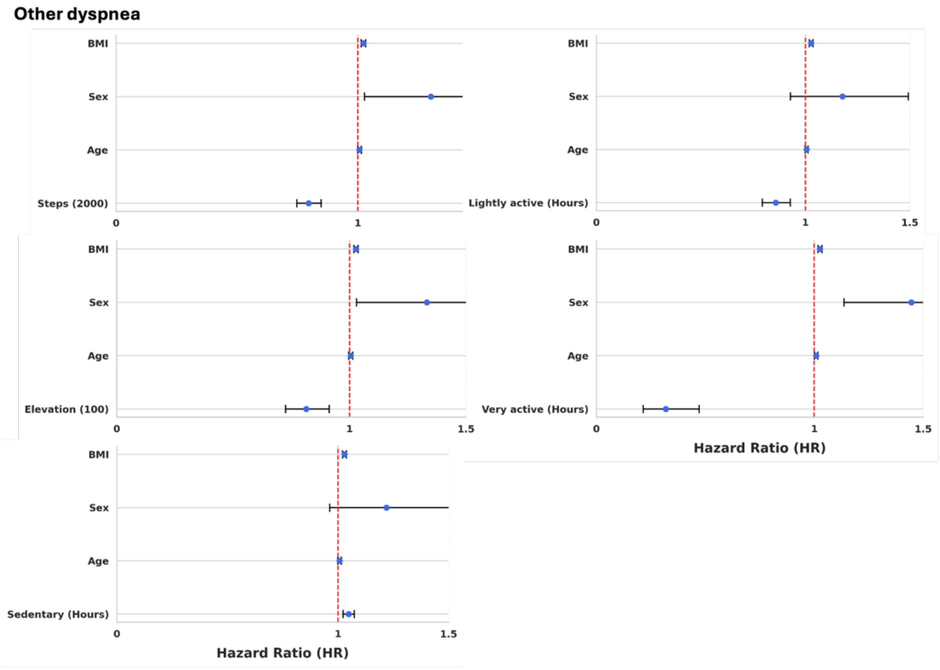


**Supplementary Fig. 6** Forest plots from Cox proportional hazard models for associations between Fitbit activity metric and other dyspnea.

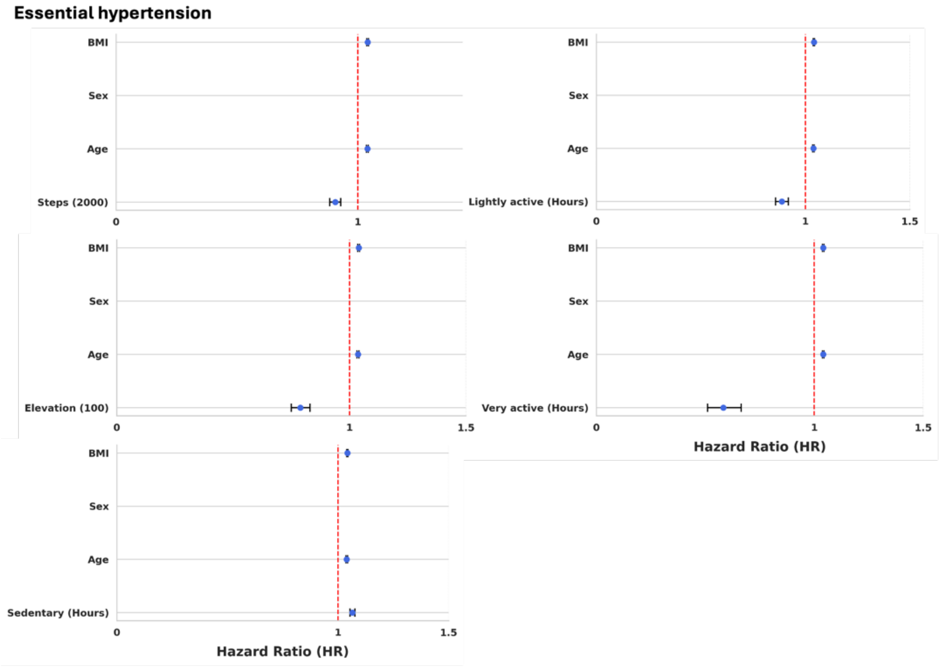


**Supplementary Fig. 7** Forest plots from Cox proportional hazard models for associations between Fitbit activity metric and essential hypertension.

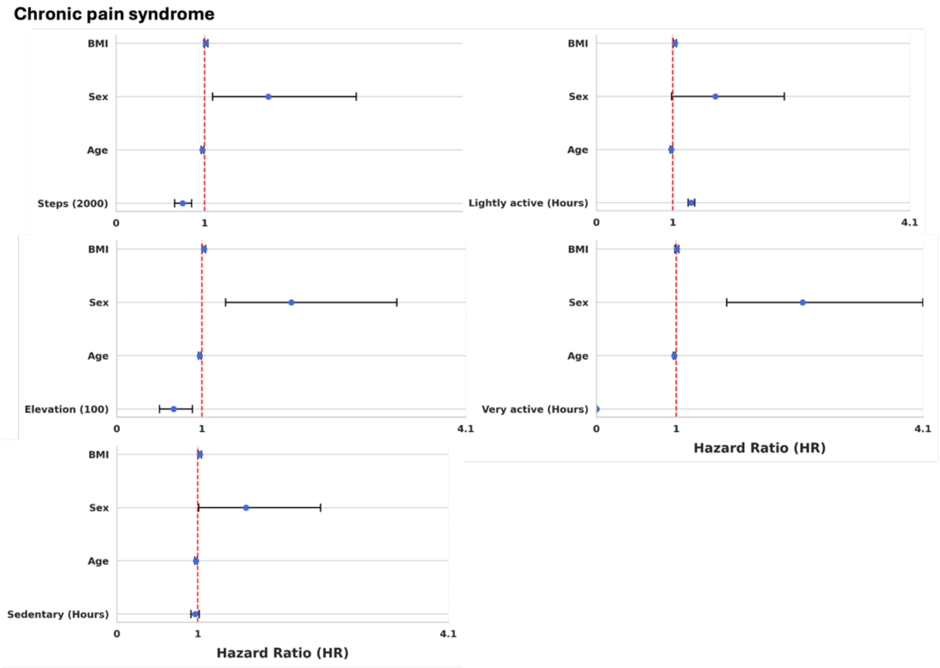


**Supplementary Fig. 8** Forest plots from Cox proportional hazard models for associations between Fitbit activity metric and chronic pain syndrome.
